# Changes of Humoral Immunity Response in SARS-CoV-2 Convalescent Patients over 8 months

**DOI:** 10.1101/2020.11.06.20227439

**Authors:** Pai Peng, Jie Hu, Hai-jun Deng, Bei-zhong Liu, Kai Wang, Ni Tang, Ai-long Huang

## Abstract

Many countries around the world have all seen a sharp rise in COVID-19 cases as the second wave since the beginning of October 2020. Decline of antibodies response to severe acute respiratory syndrome coronavirus (SARS-CoV-2) that was reported exclusively in the early month increases the risk of reinfection for convalescent individuals. There is a current need to follow the maintenance of special antibodies against SARS-CoV-2. Here, we reported changes of antibodies against SARS-CoV-2 in convalescent patients over 8 months. Antibodies of all 20 participants targeting SARS-CoV-2 spike receptor binding-domain (RBD) had decreased from a mean OD_450_ value 1.78 to 0.38 over 8 months. The neutralizing antibody (NAb) titers decreased from the mean ID_50_ value 836 to 170. The NAb titers were significantly correlated with IgG level during 8 months (*P*<0.001). Furthermore, while RBD-specific IgG existence of 25% (5/20) convalescent plasma was undetectable, the NAb titers of 15% (3/20) convalescent plasma decreased below the threshold. In addition, compared to wild-type SARS-CoV-2 (S-D614), lower titers of neutralizing antibodies against its G614 variant were shown at 8 months after symptom onset. This study has important implications when considering antibody protection against SARS-CoV-2 reinfection.

## Methods

Twenty patients who had recovered from COVID-19 were included in our cohort (see Table). Blood samples were obtained in February and October with correspond to a median of 25 days (range 5 to 33 days) and 230 days (range 221 to 248 days) after symptom onset (Figure A). Enzyme-linked immunosorbent assay (ELISA) was performed to evaluate the presence of anti-SARS-CoV-2 spike (S) receptor-binding domain (RBD) IgG over 8 months. A preliminary positive cutoff was set with the mean value of negative controls above 3 standard deviation^1^. The neutralizing antibodies (NAbs) were measured by SARS-CoV-2 wild-type (S-D614) and S-G614 mutant pseudovirus-based assays in 293T-ACE2 cells. The inhibitory dose (ID_50_) was calculated as the titers of NAbs.

## Results

Antibodies of all 20 participants targeting SARS-CoV-2 spike RBD had decreased from a mean OD_450_ value 1.78 (range 0.55 to 2.72) to 0.38 (range 0.15 to 1.01) over 8 months. When OD_450_ value was less than 0.26, the specimen was considered seronegative. At the follow-up time point 2 (in October), IgG level of 5 participants (25%) became negative (Figure B). A similar decline had been observed in the pseudovirus neutralization assay. The NAb titers decreased from the mean ID_50_ value 836.55 (range 263-1160) to 170.30 (range 33 to 365). Among them, NAb titers of 3 participants (15%) were lower than the threshold at 8 months after symptom onset (Figure C). Moreover, the NAb titers were significantly correlated with IgG levels (Figure D).

Cross-protective role of neutralizing antibodies at 8 months after symptom onset were evaluated in SARS-CoV-2 S-G614, which has been the dominant form of SARS-CoV-2 worldwide. NAb titers of 5 participants (25%) decreased below the threshold against S-G614 mutant pseudovirus. Moreover, there was a statistically significant difference in neutralizing efficacy of convalescent plasma to SARS-CoV-2 S-D614 and S-G614 mutant pseudovirus (Figure E).

## Discussion

In this study, we reported changes of humoral immunity response in SARS-CoV-2 convalescent patients over 8 months. In agreement with previous follow-up studies within a shorter time, decline of both IgG and NAb level were observed^1-3^. Furthermore, better significant correlation between IgG and NAb level in February than in October indicates that anamnestic immune response and other protective immunity should be evaluated in the context of low-level neutralizing antibodies^4^.

Facing the challenge of the second wave of SARS-CoV-2, the risk of reinfection by the currently dominant SARS-CoV-2 S-G614 variant is worthy to be considered, especially for Chinese convalescent patients infected by wild-type SARS-CoV-2.The weaker neutralizing activity to S-G614 mutant pseudovirus has been shown. In two samples, neutralizing antibody titers even quickly decrease from 1:99 or 1:122 to near the limit of detection. That might be a warning about the possible loss of protective ability of convalescent plasma with lower titers to SARS-CoV-2 S-G614 variant, like the reinfection case found in Hongkong^5^. Therefore, more data about the longevity of humoral immunity are needed to evaluate the effectiveness of herd immunity.

According to the current data, evaluation of the duration of humoral immunity response is limit. Meanwhile, there are limitations to our study on the lack of correlation analysis between clinical characteristics and specific antibody levels due to the small sample size, as well as on the lack of data to evaluate anamnestic immune response.

**Figure.**
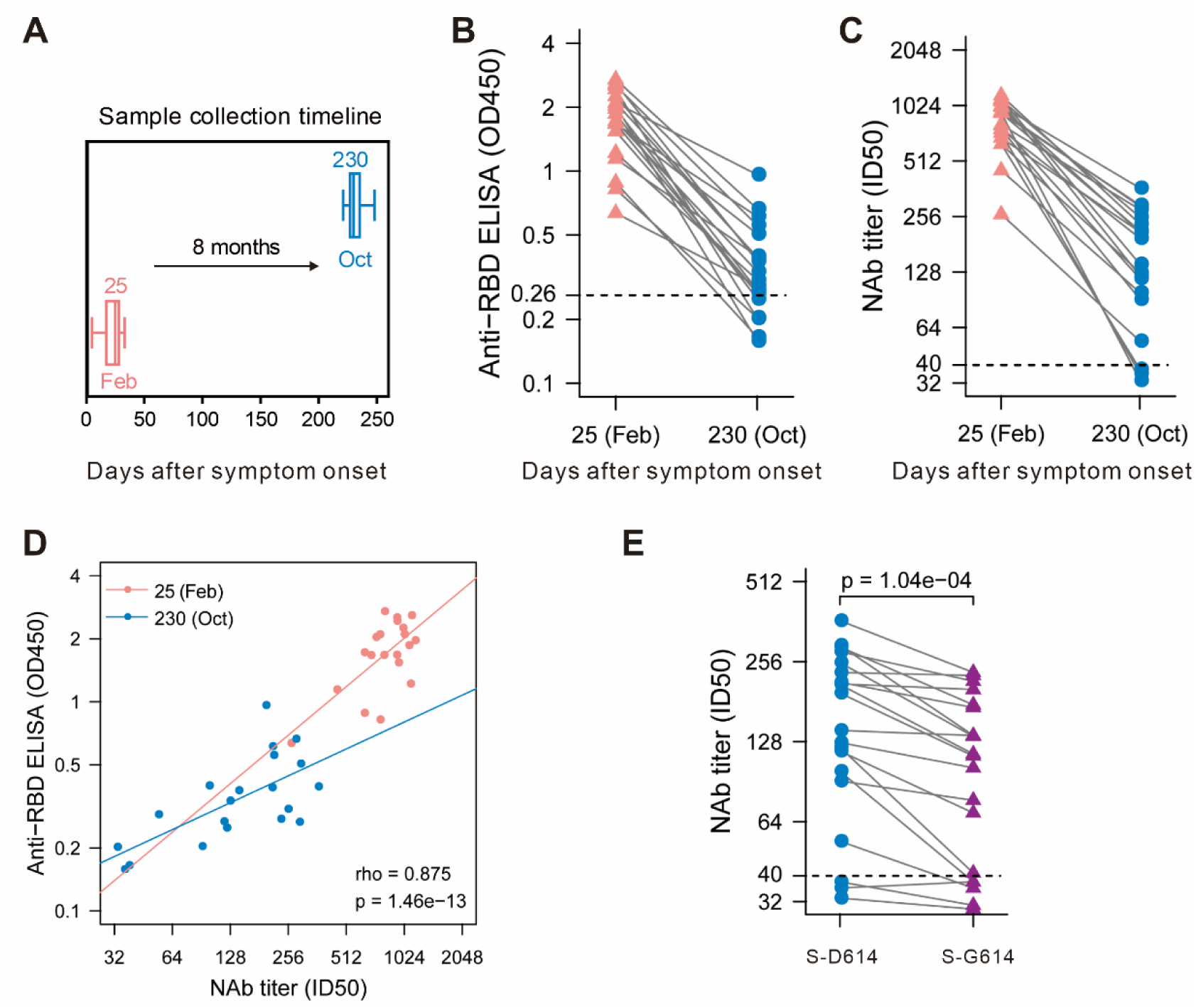
Maintenance of humoral response to SARS-CoV-2 in convalescent patients over 8 months. (**A**) Blood samples were collected in February and October. Enzyme-linked immunosorbent assay (ELISA) **(B)** and pseudovirus-based neutralization assay **(C)** were performed to detect IgG levels and neutralizing antibody (NAb) titers against SARS-CoV-2. The thresholds of detection were 0.26 of OD_450_ value and 1:40 of ID_50_, separately. **(D)** Correlation of IgG and NAb level. **(E)** Different neutralizing activity of convalescent plasma between SARS-CoV-2 S-D614 and S-G614 mutant at 8 months after symptom onset.

**Table.** Clinical characteristics of 20 convalescent individuals in the cohort^*^.

| Characteristics | Patients<br>(N=20) |
| --- | --- |
| Age (median, IQR) | 51.5<br>(44.8-65.0) |
| Sex (Male/Female) |  |
| Male | 11 (55.0%) |
| Female | 9 (45.0%) |
| Exposure |  |
| From Wuhan | 3 (15.0%) |
| Close contacts | 17 (85.0%) |
| Severity |  |
| Non-severe | 18 (90.0%) |
| Severe | 2 (10.0%) |
| Hospitalization (days) | 25 (18-28) |
| Comorbidities |  |
| Hypertension | 4 (20.0%) |
| Cardiovascular disease | 2 (10.0%) |
| Diabetes | 3 (15.0%) |
| COPD | 1 (5.0%) |
| Chronic liver disease | 1 (5.0%) |
| Signs and symptoms |  |
| Fever | 7 (35.0%) |
| Fatigue | 1 (5.0%) |
| Dry cough | 6 (30.0%) |
| Inappetence | 1 (5.0%) |
| Myalgia | 1 (5.0%) |
| Dypnea | 2 (10.0%) |
| Expectoration | 4 (20.0%) |
| Pharyngalgia | 1 (5.0%) |
| Dizziness | 1 (5.0%) |
| Vomiting | 1 (5.0%) |
| Chill | 2 (10.0%) |
| Rhinorrhea | 1 (5.0%) |
| Chest stuffiness | 1 (5.0%) |

\*Blood samples were obtained at Yongchuan Hospital of Chongqing Medical University, Chongqing, China

## Data Availability

The data supporting the findings of this study are available from the authors upon request.

## Author Contributions

PaiPeng and Jie Hu had full access to all of the data in the study and take responsibility for the integrity of the data and the accuracy of the data analysis.

*Concept and design:* Ai-long Huang, Ni Tang, Kai Wang, Pai Peng, Jie Hu.

*Acquisition, analysis, or interpretation of data:* All authors.

*Drafting of the manuscript:* Pai Peng.

*Critical revision of the manuscript for important intellectual content:* All authors.

*Statistical analysis:* Hai-jun Deng.

*Administrative, technical, or material support:* Ai-long Huang, Ni Tang, Kai Wang, Pai Peng, Jie Hu.

*Supervision:* Ai-long Huang, Ni Tang, Kai Wang.

## References

1. Ripperger TJ, Uhrlaub JL, Watanabe M, et al. Orthogonal SARS-CoV-2 Serological Assays Enable Surveillance of Low Prevalence Communities and Reveal Durable Humoral Immunity. Immunity. 2020.

2. Patel MM, Thornburg NJ, Stubblefield WB, et al. Change in antibodies to SARS-CoV-2 over 60 days among health care personnel in Nashville, Tennessee. JAMA. 2020.

3. Wang K, Long Q-X, Deng H-J, et al. Longitudinal dynamics of the neutralizing antibody response to SARS-CoV-2 infection. Clinical Infectious Diseases. 2020.

4. Chen Y, Zuiani, A., Fischinger, S., Mullur, J., Atyeo, C., Travers, M., Lelis, F.J.N. P, K.M., Martin, H., Tong, P., Gautam, A., Habibi, S., Bensko, J., Gakpo, D., Feldman, J.,, Hauser BM, Caradonna, T.M., Cai, Y., Burke, J.S., Lin, J., Lederer, J.A., Lam, E.C., Lavine, C.L.,, Seaman MS, Chen, B., Schmidt, A.G., Balazs, A.B., Lauffenburger, D.A., Alter, G., Wesemann, D.R.,. Quick COVID-19 Healers Sustain Anti-SARS-CoV-2 Antibody Production. Cell. 2020.

5. To KK-W, Hung IF-N, Ip JD, et al. COVID-19 re-infection by a phylogenetically distinct SARS-coronavirus-2 strain confirmed by whole genome sequencing. Clinical infectious diseases. 2020.

